# Evaluating the effects of cardiometabolic exposures on circulating proteins which may contribute to SARS-CoV-2 severity

**DOI:** 10.1101/2020.09.10.20191932

**Authors:** Tom G Richardson, Si Fang, Ruth E Mitchell, Michael V Holmes, George Davey Smith

## Abstract

**Background:** Developing insight into the pathogenesis of severe acute respiratory syndrome coronavirus 2 (SARS-CoV-2) is of critical importance to overcome the global pandemic caused by coronavirus disease 2019 (covid-19). In this study, we have applied Mendelian randomization (MR) to systematically evaluate the effect of 10 cardiometabolic risk factors and genetic liability to lifetime smoking on 97 circulating host proteins postulated to either interact or contribute to the maladaptive host response of SARS-CoV-2.

**Methods:** We applied the inverse variance weighted (IVW) approach and several robust MR methods in a two-sample setting to systemically estimate the genetically predicted effect of each risk factor in turn on levels of each circulating protein. Multivariable MR was conducted to simultaneously evaluate the effects of multiple risk factors on the same protein. We also applied MR using cis-regulatory variants at the genomic location responsible for encoding these proteins to estimate whether their circulating levels may influence SARS-CoV-2 severity.

**Findings:** In total, we identified evidence supporting 105 effects between risk factors and circulating proteins which were robust to multiple testing corrections and sensitivity analyses. For example, body mass index provided evidence of an effect on 23 circulating proteins with a variety of functions, such as inflammatory markers c-reactive protein (IVW Beta=0.34 per standard deviation change, 95% CI=0.26 to 0.41, P=2.19×10^−16^) and interleukin-1 receptor antagonist (IVW Beta=0.23, 95% CI=0.17 to 0.30, P=9.04×10^−12^). Further analyses using multivariable MR provided evidence that the effect of BMI on lowering immunoglobulin G, an antibody class involved in protecting the body from infection, is substantially mediated by raised triglycerides levels (IVW Beta=-0.18, 95% CI=-0.25 to -0.12, P=2.32×10^−08^, proportion mediated=44.1%). The strongest evidence that any of the circulating proteins highlighted by our initial analysis influence SARS-CoV-2 severity was identified for soluble glycoprotein 130 (odds ratio=1.81, 95% CI=1.25 to 2.62, P=0.002), a signal transductor for interleukin-6 type cytokines which are involved in the body’s inflammatory response. However, based on current case samples for severe SARS-CoV-2 we were unable to replicate findings in independent samples.

**Interpretation:** Our findings highlight several key proteins which are influenced by established exposures for disease. Future research to determine whether these circulating proteins mediate environmental effects onto risk of SARS-CoV-2 are warranted to help elucidate therapeutic strategies for covid-19 disease severity.

**Funding:** The Medical Research Council, the Wellcome Trust, the British Heart Foundation and UK Research and Innovation.

## Introduction

On the 11^th^ of March 2020 the World Health Organisation (WHO) declared the coronavirus disease 2019 (covid-19) a global pandemic^1^. Although strict lockdown measures have been enforced in many countries to control the spread of infection, the number of deaths worldwide which have been attributed to severe acute respiratory syndrome coronavirus 2 (SARS-CoV-2) continues to rise^2^. Furthermore, despite widespread ongoing biomedical research it remains unclear why some individuals develop severe symptoms of SARS-CoV-2 once contracting covid-19, whereas an estimated 80% of individuals display either asymptomatic or mild infections^3^. It is becoming increasingly evident however based on findings from the literature that established cardiometabolic disease risk factors play a role in the severity of symptoms for SARS-CoV-2^4^’^5^.

To address this critical and urgent challenge, researchers in the field, led by colleagues from the MRC Epidemiology Unit, have rapidly generated a curated dataset concerning the genetic architecture of 97 unique proteins which may be involved in influencing SARS-CoV-2 severity^6^. These include inflammatory cytokines which are involved in the body’s immune response to infection, proteins involved in fibrinolysis and blood coagulation, antibodies which play a critical role in the body’s immune response to infection (such as immunoglobulin G) and gene products which have been reported to interact with SARS-CoV-2 proteins in human cells^7^. A complete list of these proteins can be found in **Supplementary Table 1**.

This curated resource provides an opportunity to undertake Mendelian randomization (MR) analyses to develop insight into the environmental risk factors that influence these SARS-CoV-2-related proteins, as well as potential downstream consequences on risk of covid-19. MR can be implemented as a form of instrumental variable analysis which exploits the random assortment of genetic alleles at birth under Mendel’s laws of Inheritance^8^’^9^. As such genetic variants can be leveraged as instrumental variables to investigate causal relationships between conventional exposures (such as cardiometabolic risk factors) and outcomes (such as circulating proteins) (Figure 1A). As these inherited genetic variants are fixed at conception, MR is typically robust to confounding factors and reverse causation which can bias analyses in an observational setting which do not make use of human genetics data.

**Figure 1:**
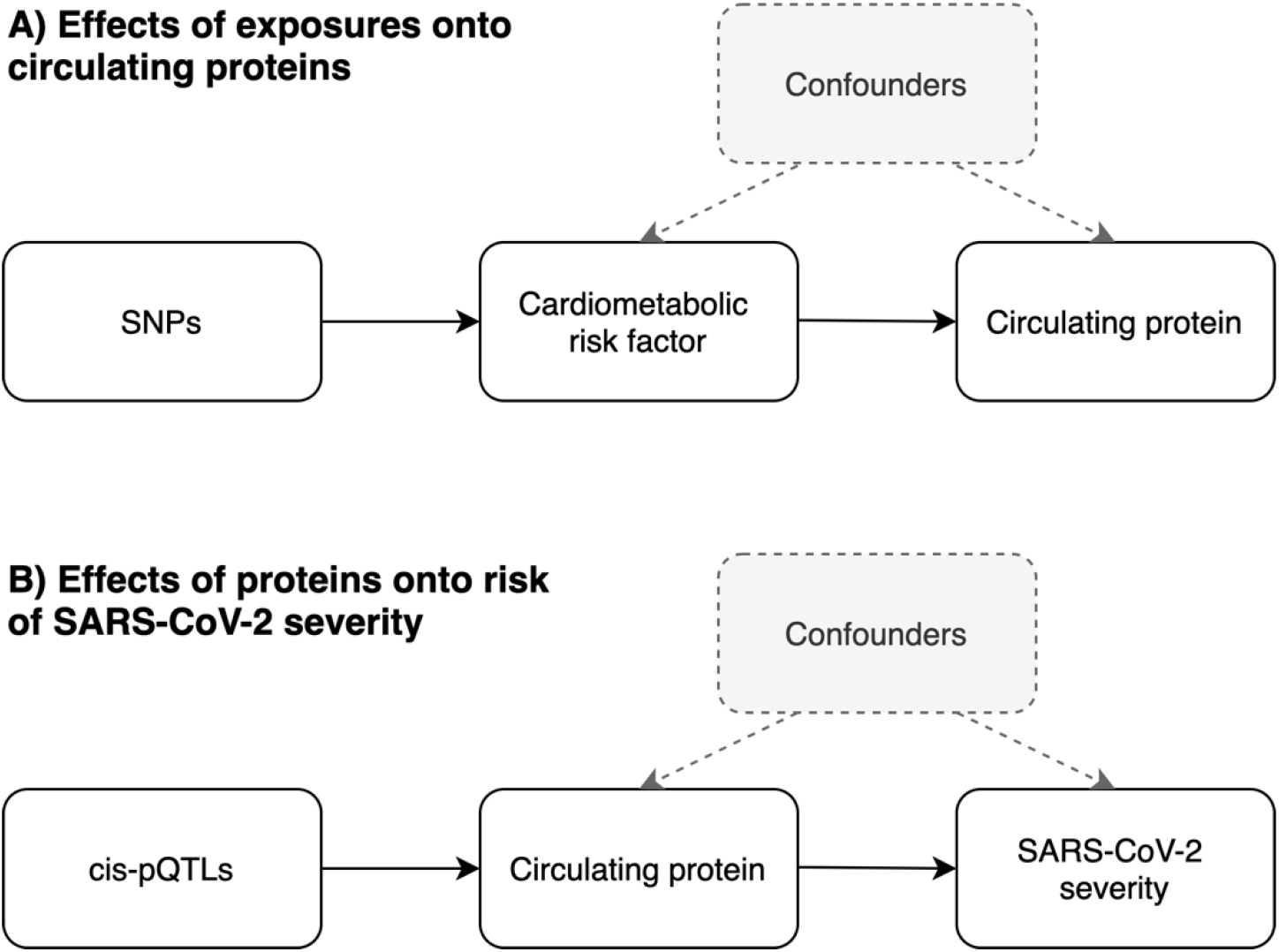
Directed acyclic graphs (DAGs) to illustrate the analysis undertaken in this study using Mendelian randomization. A) We firstly leveraged genetic variants (referred to as single nucleotide polymorphisms (SNPs)) to systematically estimate the effect of 11 risk factors on 97 circulating proteins related to SARS-CoV-2. B) For proteins highlighted in the initial analysis, we applied MR to estimate their genetically predicted effects on risk of SARS-CoV-2 severity. Instruments for proteins were SNPs robustly associated with their levels and located in the genome at the encoding genes region (commonly referred to as cis-protein quantitative trait loci (cis-pQTL)).

In this study we systematically applied MR to estimate the effects of 10 cardiometabolic exposures and genetic liability to lifetime smoking in turn on each of the SARS-CoV-2 prioritised proteins. This was followed by a series of sensitivity analyses as well as applying multivariable MR to evaluate whether exposures independently influence the same circulating protein or act along overlapping causal pathways. We also sought to investigate the potential effects of proteins highlighted by this analysis on risk of severe covid-19 using data from recently conducted genome-wide association studies (GWAS).

## Methods

### Data resources

#### Deriving genetic instruments for modifiable exposures

We obtained genetic instruments for 11 exposures using data from large-scale GWAS. These were body mass index (BMI), systolic blood pressure (SBP), diastolic blood pressure (DBP), high density lipoprotein (HDL) cholesterol, low density lipoprotein (LDL) cholesterol, triglycerides, apolipoprotein A-I (Apo A-I), apolipoprotein B (Apo B), genetic liability to lifetime smoking, waist-hip-ratio adjusted for BMI and childhood adiposity based on reported body size at age 10^10-13^. Details on the study characteristics for the GWAS used to derive these instruments can be found in **Supplementary Table 2**.

We undertook linkage disequilibrium (LD) clumping to identify independent genetic instruments for these 11 exposures assessed using the software PLINK^14^. This process involves removing genetic variants which are correlated with the mostly strongly associated variant with a trait of interested in a region based on pairwise LD (using r^2^ < 0.001 in this study) using a reference panel of 503 individuals of European descent from phase 3 (version 5) of the 1000 genomes project^15^.

#### Quantitative trait loci data for SARS-CoV-2-reIated proteins

All pQTL summary statistics for 97 unique proteins were obtained from the https://omicscience.org/apps/covidpgwas Webserver^6^. Details on how these pQTL were derived are described in detail in the study by Pietzner et al and outlined in **Supplementary Figure** 1. Briefly, plasma samples from 10,708 individuals from the Fenland population-based cohort study were eligible for analysis after exclusions. In total, 409 circulating proteins which were prioritised due to any of the following criteria; evidence suggesting that they interact with SARS-CoV-2 (n=332)^7^, associated with disease severity (n=26)^16^, involved in viral entry (n=2)^17^ or that they are clinical biomarkers of adverse, prognosis, complications and disease deterioration (n=54)^18-21^. Of the proteins, SOMAscan proteomic assays were used to derive data on 179 of them.

In the same set of participants, imputed genotype data on 17,652,797 genetic variants were available after imputation using the UK10K+1000G phase3 reference panel. Summary statistics were available in a total of 97 unique proteins from the Webserver as they had at least 1 pQTL acting *in cis*, which is defined here as genetic variants robustly associated with circulating proteins (based on P<5×10^−08^) and located within a 1Mb window around the genes responsible for encoding them.

We undertook LD clumping as before to identify pQTL to be used as instruments in MR analyses. However, for protein instrumental variables we applied a more lenient LD threshold of r^2^<0.2 to identify weakly correlated pQTL all within a cis-window of 1Mb either side of the lead cis-pQTL for each protein analysed. MR was then undertaken for protein targets whilst taking into account LD structure of pQTL as proposed previously^22^.

#### Covid-19 GWAS datasets

Genetic estimates on SARS-CoV-2 were obtained using data from a GWAS of severe covid-19^23^ based on 1980 patients from intensive care units and wards at seven hospital located in the pandemic epicenters in Italy and Spain. Severe covid-19 was defined as hospitalization with respiratory failure and a confirmed SARS-CoV-2 viral RNA polymerase-chain-reaction (PCR) test using nasopharyngeal swabs or other biologic fluids. Effect estimates from these GWAS were mapped to hg19 coordinates using the LiftOver tool (https://genome.sph.umich.edu/wiki/LiftOver). We analysed GWAS data on severe SARS-CoV-2 to data based on reported covid-19 symptoms can potentially lead to misleading conclusions^24^. We also sought out replication of findings using GWAS results from the covid-19 host genetic initiative^25^ using data on hospitalized covid-19 cases compared to population controls (https://www.covidl9hg.org/results/), as well as a GWAS of mortality attributed to covid-19 in the UK Biobank study compared to population controls based on analyses by Johnson and colleagues^26^ (available at https://grasp.nhlbi.nih.gov/Covidl9GWASResults.aspx).

### Statistical analysis

We firstly applied MR to estimate the effect of each of the 11 exposures in turn on each protein in a two-sample setting^27^. Initially we used the inverse variance weighted (IVW) approach which takes the SNP-outcome estimates and regresses them on those for the SNP-exposure associations. This provides an overall weighted estimate of the causal effect which is based on the inverse of the square of the standard error for the SNP-outcome association^28^. We applied a correction using false discovery rate (FDR)<5% to these results to account for multiple testing. This threshold has been used in this study as a heuristic to highlight findings with the strongest statistical evidence to investigate in further detail.

For effects which survived FDR corrections we applied other MR methods as sensitivity analyses. This firstly involved applying the weighted median and MR-Egger approaches which are more robust to horizontal pleiotropy in comparison to the IVW method^29,30^. We also applied the MR directionality test (also referred to as the ‘Steiger method’) to support evidence that our genetic variant is a valid instrument for the exposure in line with the underlying assumptions of MR^31^. Multivariable MR was undertaken to evaluate the direct effects of exposures on circulating proteins whilst accounting for the effects of other exposures^32,33^. In doing so, we were able to investigate whether the effect of exposures on proteins were putatively mediated via another exposure. To estimate the proportion mediated we applied the product method described previously by Burgess and colleagues^34^ using genetic instruments from the GIANT consortium for BMI to avoid overlapping samples with the UK Biobank^35^.

We next applied MR to investigate the genetically predicted effects of circulating proteins robust to FDR corrections and sensitivity analyses in the previous analysis on severe SARS-CoV-2 using GWAS results from Ellinghaus et al. This was undertaken by applying the IVW method which uses correlated cis-regulatory variants whilst accounting for their correlation structure^36^. For proteins which provided strong evidence of an effect using the IVW method, we also applied the MR-Egger approach whilst accounting for correlation structure amongst instruments. We also attempted to replicate findings using the GWAS data from the covid-19 HGI and Johnson et al analyses^25,26^. Finally, we applied the cis-correlated IVW approach systematically to all proteins with at least 2 cis-pQTL (as this is the minimum number required for the IVW method) for each covid-19 GWAS dataset. This allowed us to highlight proteins which may play a role in disease but are not strongly under the influence of modifiable risk factors.

All analyses were undertaken using the ‘TwoSampleMR’ and ‘MendelianRandomization’ packages using R (version 3.5.1)^37,38^. The forest plot in Figure 2 was generated using ‘ggplot2’ v2.2.1^39^. Figure 3 was generated using the LD link resource^40^.

**Figure 2:**
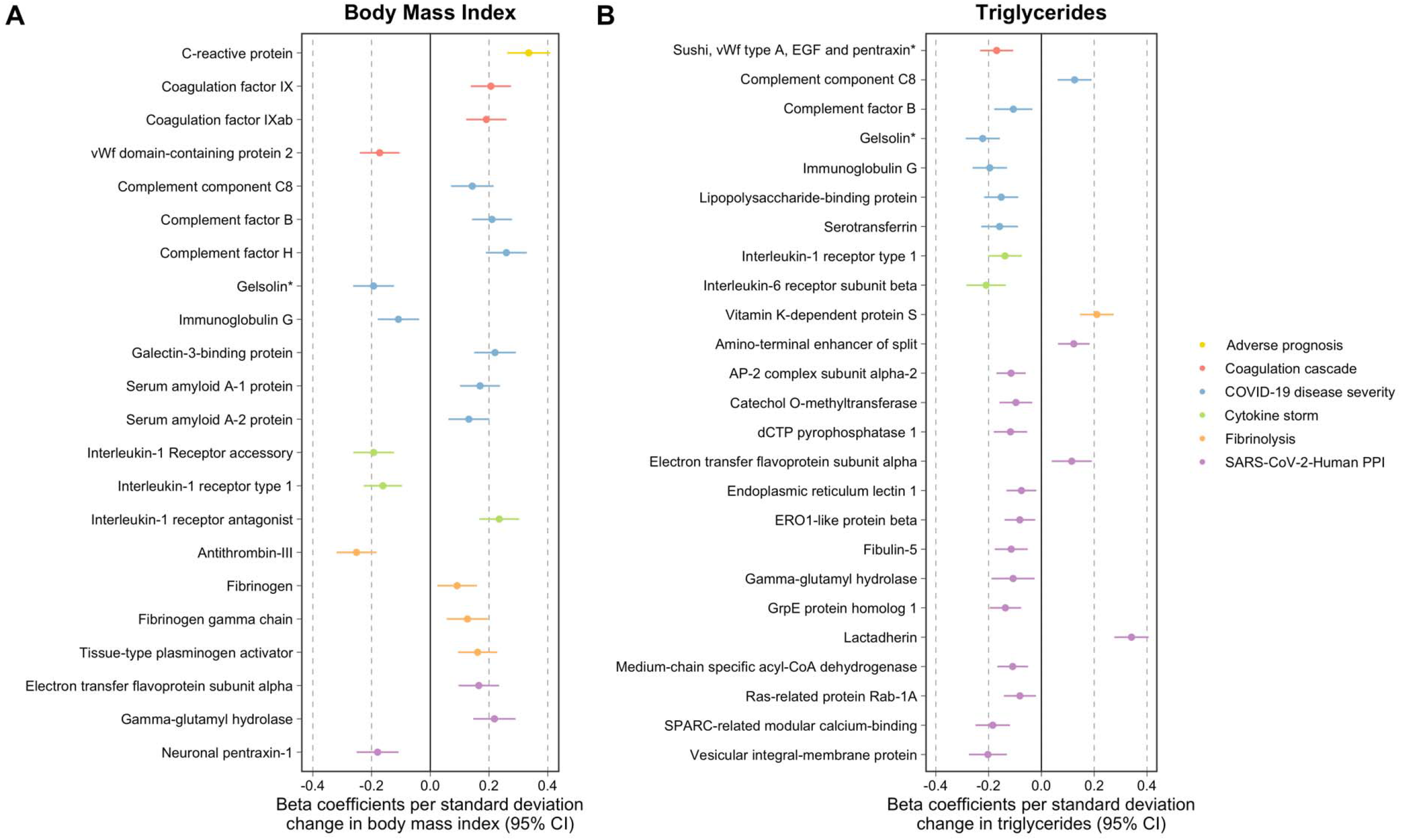
Forest plots illustrating the Mendelian randomization estimates of genetically predicted A) body mass index and B) triglycerides on circulating proteins related to SARS-CoV-2 severity. Proteins have been grouped and coloured based on their subcategories. * Gelsolin corresponds to soma ID 16607-78 as a separate epitope is also available on the SomaLogic assay for this protein.

## Results

### A systematic Mendelian randomization analysis of circulating proteins

Across the 11 exposures assessed, there were 253 genetically predicted effects on circulating proteins which survived FDR<5% corrections using the IVW method **(Supplementary Table 3)**. Amongst top findings was a strong effect of BMI on C-reactive protein (CRP) levels (Beta=0.34 per standard deviation change in BMI, 95% CI=0.26 to 0.41, P=2.19×10^−16^) which is a well-established marker of chronic inflammation^41^. Elsewhere, there was strong evidence of an effect of HDL cholesterol on elevated levels of serum amyloid A-1 (Beta=0.23, 95% CI=0.16 to 0.29, P=1.59×10^−12^) and A-2 (Beta=0.24, 95% CI=0.17 to 0.30, P=4.38×10^−13^) proteins. Undertaking sensitivity analyses found 106 effects that were robust to FDR<5% corrections using either the weighted median or MR-Egger methods **(Supplementary Tables 4 & 5)**. The MR directionality tests provided evidence that assumptions regarding directionality may have been violated for one of these effects, which was between waist-hip-ratio adjusted for BMI and the protein ITIH3 (IVW Beta=-0.31, 95% CI=-0.46 to -0.16, P=4.07×10^−05^) **(Supplementary Table 6)**. IVW estimates for the 105 effects which were robust to sensitivity analyses can be found in **Supplementary Table 7**. An overview of the analytical pipeline applied in this section including method and datasets used can be found in **Supplementary Figure 2**.

Amongst the exposures which contributed most to the remaining 105 effects were BMI (23 effects) and triglycerides (27 effects). As illustrated in **Figure 2**, the effects driven by BMI were typically spread across the 6 subcategories of circulating proteins. This included effects on coagulation factor IX (IVW Beta=0.21, 95% CI=0.14 to 0.27, P=2.42×10^−07^), tissue-type plasminogen activator (Beta=0.16, 95% CI=0.09 to 0.23, P=2.38×10^−06^) and cytokines including interleukin-1 receptor antagonist (Beta=0.23, 95% CI=0.17 to 0.30, P=9.04×10^−12^). In contrast, the majority of triglycerides effects were found to be on proteins allocated to the SARS-CoV-Human protein-protein interaction (PPI) or covid-19 disease severity subcategories. There were exceptions to this however, such as an effect on clotting factor vitamin K-dependent protein S (Beta=0.21, 95% CI=0.15 to 0.27, P=1.25×10^−10^) and cytokine signal transducer interleukin-6 receptor subunit beta (Beta=-0.21, 95% CI=-0.28 to -0.13, P=4.12×10^−08^). We also note the conflicting directions of effect which risk factors have on the proteins assessed even within the same category. For example, within the fibrinolysis category BMI provided evidence of an effect on higher levels of fibrinogen (Beta=0.09, 95% CI=0.02 to 0.16, P=0.008), as well as an inverse effect on antithrombin III (Beta=-0.25, 95% CI=-0.32 to -0.18, P=4.43×10^−13^). Findings from the literature supported the direction of effect between BMI and circulating proteins for various effects identified in this analysis (CRP, Factor B and H, the interleukin 1 family of proteins, SAA/2, fibrinogen and antithrombin III), although for others there was no clear prior evidence suggesting that obesity influences their levels **(Supplementary Table 8)**.

BMI is recognized to causally influence triglycerides and we therefore undertook multivariable MR to evaluate the direct effects of BMI and triglycerides on the 7 proteins which they had in common based on univariable estimates in the previous analysis. The majority of these effects remained robust after accounting for the effects of the other exposure, suggesting that BMI and triglycerides influence these proteins directly via separate causal pathways **(Supplementary Table 9)**. The notable exception to this was the effect of BMI on immunoglobulin G, an antibody class involved in the body’s immune response to infection. Effect estimates for BMI on this circulating protein identified in the univariable analysis (Beta=-0.11, 95% CI=-0.18 to 0.04, P=0.02) attenuated to the null when accounting for the effect of triglycerides (Beta=-0.06, 95% CI=-0.13 to 0.02, P=0.13). This suggests that BMI indirectly lowers immunoglobulin G due to its influence on raising triglyceride levels. We estimated that 44% of the BMI effect on immunoglobulin G was mediated via triglycerides using mediation MR.

### Harnessing cis-regulatory variants to evaluate effects of circulating proteins on risk of severe SARS-CoV-2

For each protein highlighted in the previous analysis, we undertook MR using cis-pQTL as instruments to estimate their effects on risk of severe SARS-CoV2-2 **(Supplementary Table 10)**. The protein which provided the strongest evidence that it may influence risk of covid-19 was gp130, soluble, also known as glycoprotein 130 (odds ratio (OR)=1.81 increased risk of severe SARS-CoV2-2 per 1-fold increase in gp130, 95% CI=1.25 to 2.62, P=0.002). This protein is encoded by the *IL6ST* gene and is responsible for signal transduction with all members of the interleukin 6 receptor family^42^. There were 18 weakly correlated pQTL scattered across this locus used as instrumental variables as illustrated in **Figure 3**. Their pairwise LD correlations can be found in **Supplementary Table 11**. Although another cytokine gene is also located in this region (*IL31RA*), single variant associations for these 18 pQTL with severe SARS-CoV-2 suggested that this alternate target is unlikely to be responsible for this putative effect **(Supplementary Table 12)**. This is because the two lead pQTLs largely responsible for driving the overall IVW estimate were rs929108 (P=0.002), which is located downstream of *IL6ST* (i.e. the opposite side compared to *IL31RA*) and rs6875155 (P=0.006), located within the gene body of *IL6ST* itself.

**Figure 3:**
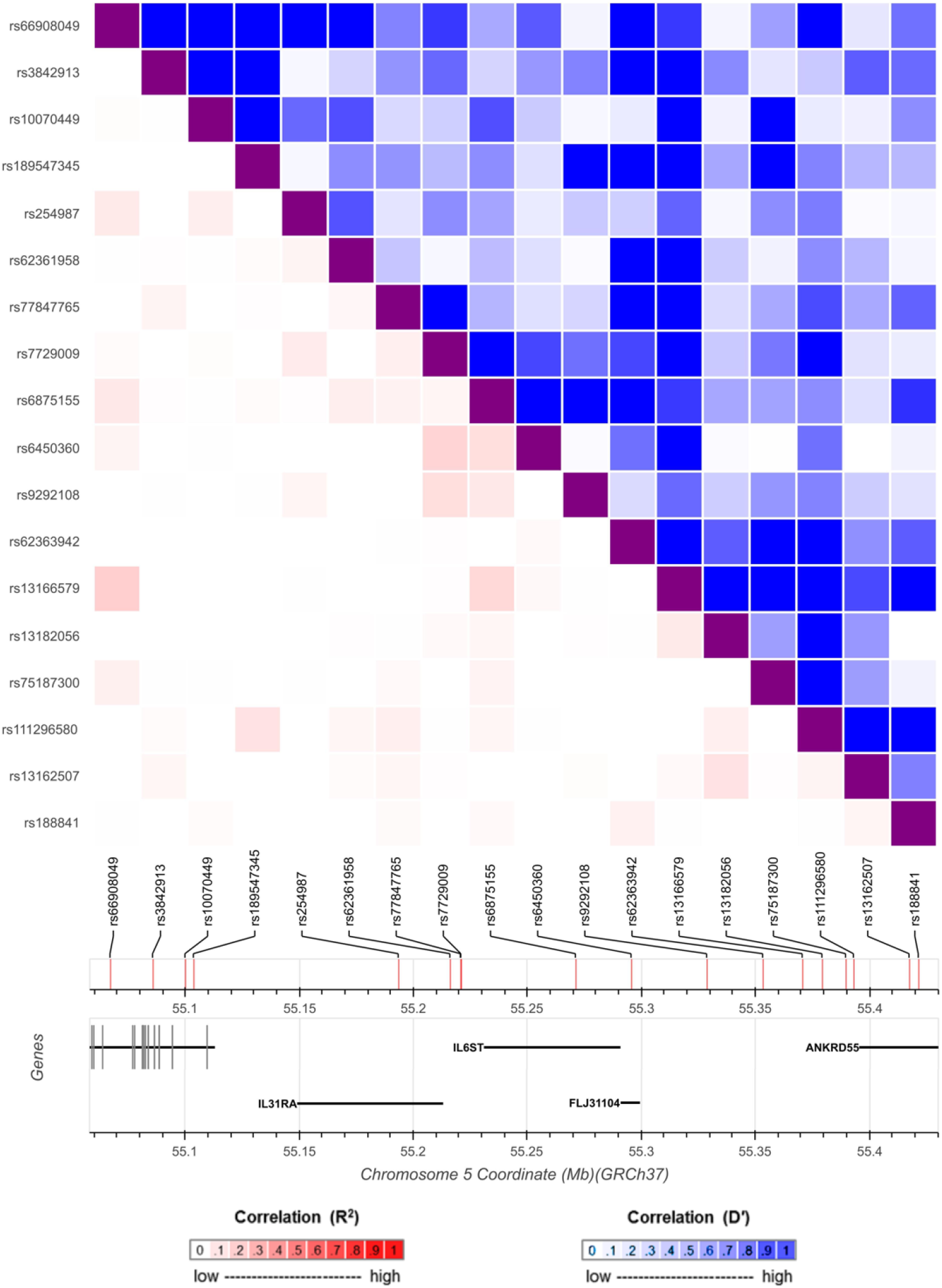
A diagram of genetic locus used to identify protein quantitative trait loci (pQTL) as instrumental variables in the Mendelian randomization analysis for glycoprotein 130 (encoded by IL6ST) and SARS-CoV-2 severity. The heatmap represents the linkage disequilibrium structure at this region amongst the pQTL used with the bottom-left section representing pairwise r^2^ coefficients (red) and the upper-right section illustrating pairwise D’ values (blue).

Applying the MR-Egger method accounting for correlation structure using the 18 *IL6ST* instruments provided effect estimates which included the null (OR=1.55, 95% CI=1.00 to 2.39, P=0.05). Furthermore, we were unable to detect robust evidence of replication using the two other covid-19 GWAS datasets (covid-19 host genetics initiative: OR=1.11, 95% CI=0.83 to 1.48, P=0.48 & Risk of death due to covid-19 death in UK Biobank: OR=1.26, 95% CI=0.79 to 2.00, P=0.34). Lastly, we applied the IVW method accounting for correlation structure to all proteins with at least 2 cis-pQTL to evaluate their genetically predicted effects on risk of covid-19. However, using current sample sizes we did not detect strong evidence that these circulating proteins influence risk of severe covid-19 based on multiple testing corrections **(Supplementary Table 13, Supplementary Table 14 & Supplementary Table 15)**.

## Discussion

We have undertaken a comprehensive Mendelian randomization study to systematically evaluate the effect of 11 established risk factors for disease on circulating levels of proteins related to SARS-CoV-2. Our main findings are that among the modifiable risk factors assessed, BMI and triglycerides showed the widest repertoire of causal effects on circulating proteins (providing evidence of causation for 23 and 27 effects, respectively). Furthermore, of the circulating proteins investigated by our study, the strongest evidence of an effect on developing severe covid-19 was identified for glycoprotein 130, which is involved in the transmission of molecular signals for inflammatory interleukin cytokines.

Amongst the 105 effects which were robust to multiple testing and sensitivity analyses there were several well-established relationships based on the literature. For example, having a high BMI is a known driver of systemic inflammation as indexed by C-reactive protein levels^41^ and acute inflammatory markers such as fibrinogen^43^. Other findings fit with the known biology of cardiometabolic risk factors and proteins identified by our analysis, such as the effect of HDL cholesterol on serum amyloid A-1 and A-2 proteins, which have previously been proposed as clinically applicable surrogates of HDL vascular functionality ^44^ Whilst our results are therefore of immediate importance for SARS-CoV-2 research, they may also be valuable for future endeavours interested in the therapeutic potential of these proteins with respect to a wide range of disease outcomes.

There were several results from our study which may assist in unravelling the complex pathogenesis of SARS-CoV-2 severity. For example, immunoglobulin G (IgG) is a class of antibodies produced by plasma B cells in the immune system in response to a pathogen ^45^. Our results indicate that having a high BMI may reduce levels of circulating IgG, suggesting that people with obesity have less of this class of antibody to help protect their body’s immune system from infection. That being said, an important consideration when interpreting this finding is that IgG levels were measured in individual’s in a healthy state and can therefore only act a proxy for IgG response to infection. Additionally, generic IgG levels were measured rather than the specific adaptive immune response to SARS-CoV-2.

Additionally, our multivariable MR estimates for the effect BMI on IgG attenuated to include the null when accounting for the effect of triglycerides on this class of antibodies. This suggests that triglycerides may mediate the lowering effect of BMI on IgG, which we estimated as 41.1% of the total effect of BMI on IgG levels being mediated via triglycerides. Further research into the role of IgG and B cell immunity in the body’s immune response to the covid-19 pathogen is therefore warranted, particularly given that IgG is being measured by tests for antibody responses to SARS-CoV-2^46^. Along with evaluating the effect of modifiable risk factors on antibody mediated immunity to covid-19, it will be critical to develop insight into how these factors influence cell mediated immunity given the emerging importance of the adaptive immune response to SARS-CoV-2^47^.

Using MR to estimate the genetically predicted effects of circulating proteins on severe covid-19 risk highlighted glycoprotein 130 as the protein with the strongest evidence of an effect on SARS-CoV-2 severity (OR=1.81, 95% CI=1.25 to 2.62, P=0.002), however we were unable to replicate these findings in larger samples. Glycoprotein 130 is encoded by the *IL6ST* gene and belongs to the interleukin-6 family of cytokines^42^. It’s activation is dependent upon the binding of cytokines with their receptors, such as interleukin-6 (IL6) with interleukin-6 receptor (IL6R)^48^. This is noteworthy due to the interest in repurposing IL6R blockers as a potential therapeutic strategy for SARS-CoV-2 ^49^. As lowering the levels of circulating IL6R will lead to lower activation of glycoprotein 130, estimates in this study suggest that this might result in reduced risk of severe SARS-CoV-2 symptoms. These findings therefore corroborate results from a recent MR study which used human genetic data to support the efficacy of IL6R inhibition as a potential treatment option for severe SARS-CoV-2 symptoms^50^. There is however conflicting evidence emerging from clinical trials evaluating the repurposing of IL6R blockers for covid-19 therapy with some already being halted^51^.

This study has several limitations which should be taken into account when interpreting its findings. The current sample sizes of the SARS-CoV-2 GWAS are (as one would expect) relatively modest compared to large-scale GWAS data which MR studies are contemporaneously applied to, meaning that our cis-pQTL analysis is likely underpowered. We analysed severe covid-19 as an outcome to mitigate reported selection bias of cases^24^ so larger sample sizes of severe SARS-CoV-2 GWAS in the future should improve the statistical power of our approach. Furthermore, although the protein GWAS data is of unprecedented sample size compared to previous large-scale pQTL analyses (n=10,708), it remains comparably modest to the sample sizes of GWAS used to derive instrument for the cardiometabolic exposures in this work. This is exaggerated by the fact that protein MRs are typically conducted using a monogenic instrument and thus genetic instruments are likely to explain a lower proportion of variance in the exposure^52^. Therefore, although we have undertaken thorough evaluations to interrogate bi-directional relationships between the exposures and proteins in this study, the discrepancies between the samples sizes makes the direction of effect challenging to orient (the majority of exposure instruments were derived using sample sizes of n=~440,000). Finally, although data from plasma pQTL studies provide an exceptional opportunity to leverage instruments for MR studies, it should be noted that serum plasma may not capture signatures confined to disease or cell-type relevant tissues. This is particularly important for a disease with a large autoimmune component such as covid-19 and further emphasis should therefore be noted when interpreting the results of our study on proteins such as IgG.

In conclusion, our MR study identified many effects between conventional risk factors and circulating proteins which provides a platform for prospective endeavours to dissect related disease pathways. Future research into the pathogenesis of the proteins highlighted by this study are warranted to discern whether they may hold therapeutic potential for covid-19 severity.

## Data Availability

All data analysed in this study is publicly accessible through the referenced web resources.

## Acknowledgements

We are extremely grateful to Maik Pietzner, Claudia Langenberg and all their colleagues who were involved in identifying the pQTL for the SARS-CoV-2-related proteins analysed in this study. We are also enormously thankful to all the authors of the Ellinghaus et al GWAS, the covid-19 Host Genetics Initiative and Dr Andrew Johnson and colleagues for sharing their summary statistics which made analyses of covid-19 risk and cause of death in this study possible.

This work was supported by the MRC Integrative Epidemiology Unit which receives funding from the UK Medical Research Council and the University of Bristol (MC_UU_00011/1). GDS conducts research at the NIHR Biomedical Research Centre at the University Hospitals Bristol NHS Foundation Trust and the University of Bristol. The views expressed in this publication are those of the author(s) and not necessarily those of the NHS, the National Institute for Health Research or the Department of Health. SF is supported by a Wellcome Trust PhD studentship in Molecular, Genetic and Lifecourse Epidemiology [108902/Z/15/Z]. TGR is a UKRI Innovation Research Fellow (MR/S003886/1). MVH works in a unit that receives funding from the UK Medical Research Council and is supported by a British Heart Foundation Intermediate Clinical Research Fellowship (FS/18/23/33512) and the National Institute for Health Research Oxford Biomedical Research Centre.

## Materials and Correspondence

This publication is the work of the authors and TGR will serve as guarantor for the contents of this paper.

